# Kidney transcriptomics signature of prospective rapid diabetic kidney disease progression

**DOI:** 10.1101/2024.07.03.24309831

**Authors:** Dianne Acoba, Anna Levin, Anna Witasp, Kerstin Ebefors, Johan Mölne, Peter J. Greasley, Jenny Nyström, Annika Wernerson, Anna Reznichenko

**Affiliations:** Translational Science & Experimental Medicine, Research and Early Development, Cardiovascular, Renal and Metabolism, BioPharmaceuticals R&D, AstraZeneca, Gothenburg, Sweden; Institut Necker Enfants Malades (INEM), Institut National de la Santé et de la Recherche Médicale (INSERM) U1151, Université Paris Cité, Paris, France; Department of Clinical Science, Intervention and Technology, Division of Renal Medicine, Karolinska Institutet, Stockholm, Sweden; Department of Physiology, Institute of Neuroscience and Physiology, Sahlgrenska Academy, University of Gothenburg, Gothenburg, Sweden; Department of Laboratory Medicine, Institute of Biomedicine, Sahlgrenska Academy, University of Gothenburg, Gothenburg, Sweden; Early Clinical Development, Research and Early Development, Cardiovascular, Renal and Metabolism, BioPharmaceuticals R&D, AstraZeneca, Gothenburg, Sweden

**Keywords:** chronic kidney disease, diabetic kidney disease, rapid progression, eGFR slope, gene expression, longitudinal study

## Abstract

Previous cross-sectional transcriptomic studies on diabetic kidney disease (DKD) kidney tissue have shown correlations between gene expression and both disease status and kidney function at time of biopsy, but longitudinal data is scarce. We utilized clinical follow-up data up to five years post-biopsy, linking the transcriptomes of diagnostic kidney biopsies to progression rates and outcomes in 19 DKD patients. Patients were stratified into “rapid progressors” and “non-rapid progressors” based on clinical parameters (eGFR slope, CKD stage advancement, degree of albuminuria, composite of kidney failure or 40% eGFR decline). Differential expression and pathway enrichment analyses were performed to identify dysregulated genes and pathways associated with rapid progression. We identified 265 genes in glomeruli and tubulointerstitium that were significantly modulated in rapid vs non-rapid DKD progression. Rapid progression-associated genes showed enrichment for well-established (extracellular matrix organization, inflammation) as well as novel pathways in the context of DKD (circadian rhythm, cytoskeleton reorganization, *NOTCH* signaling). This study illuminates kidney gene expression patterns that may be predictive of rapid progression in DKD and are distinct from those associated with cross-sectional kidney function.

## INTRODUCTION

Diabetic kidney disease is the leading cause of chronic kidney disease (CKD) and kidney failure,^1^ affecting patients with Diabetes Mellitus (DM). The etiology is multifactorial and its clinical presentation is highly heterogeneous with some patients developing DKD at an early stage of DM, while others never do despite multiple risk factors.^2^ This complexity in DKD pathogenesis is also mirrored in its progression pattern where there is a high inter-individual variability in the rate of kidney function decline and levels of albuminuria over time.^3^ Understanding the pathophysiological drivers and mechanisms of DKD progression is therefore essential to achieve early identification of patients at high risk for rapid kidney function decline as well as to enable identification of novel therapeutic targets to delay or even reverse the disease.

Over the past twenty years, numerous kidney transcriptomics studies have been made, showing significant strides in mapping the human DKD molecular landscape. These studies predominantly compared gene expression profiles of DKD and control kidneys, sometimes employing tissue microdissection, and identified a plethora of disease-implicated pathways. Such were, for example, pathways linked to diminished glomerular tissue repair,^4^ and SRGAP2a as a key gene associated with proteinuria and estimated glomerular filtration rate (eGFR).^5^ The European Renal cDNA Bank (ERCB) study demonstrated association between tubulointerstitial expression of NF-κB inflammatory pathway genes^6^ and TRAIL^7^ in DKD. In addition, the Wnt signaling pathway was found activated in DKD,^8^ and transcripts involved in inflammation, metabolism, migration, and mitochondrial dysfunction pathways showed correlations with tubulointerstitial damage.^9^ A study by Berthier et al showed that the Janus kinase-STAT pathway is up-regulated in both glomerular and tubulointerstitial compartments,^10^ while Woroniecka et al found RhoA, Cdc42, integrin and VEGF signaling to be differentially regulated in the glomeruli and inflammation-related pathways in the tubulointerstitium.^11^

The introduction of the more unbiased and high-resolution RNA sequencing (RNA-seq) methods has allowed further fine-tuning of the previous findings that were based on microarray technologies. So far, Fan et al used RNA-seq profiling of non-microdissected kidney biopsies from advanced vs early DKD patients and identified retinoic acid pathway to be renoprotective,^12^ whereas our group generated RNA-seq data on microdissected DKD kidney biopsies and reported compartment-specific changes, including glomerular upregulation of pathways associated with extracellular matrix reorganization and inflammation, and tubulointerstitial enrichment of apoptotic pathways.^13^

Together, these systematic studies on transcriptome changes in the affected kidney amassed a pool of disease-associated transcripts with hypothesized roles in DKD pathogenesis. However, as all of them used cross-sectional case-control design, it remains unknown whether the associations go beyond baseline, inferring rapid kidney function decline and progression. Thus, longitudinal studies linking baseline kidney biopsy transcriptomics data to prospectively collected clinical data on disease progression would be the next imperative step towards unravelling the molecular drivers of DKD course heterogeneity.

To leverage our previous DKD characterization study,^13^ we collected post-biopsy prospective longitudinal clinical data and stratified DKD patients based on clinically relevant measures and outcomes into rapid- and non-rapid progressors. In this way we were able to uncover rapid DKD progression gene expression signatures in each of the glomerular and tubulointerstitial compartments. Previously reported genes but also unexplored genes in the context of DKD pathophysiology were identified, thus highlighting possible novel DKD progression drivers as well as corroborating prior hypotheses.

## METHODS

### Study cohort

Patients’ baseline characteristics as well as the transcriptomics data generation from kidney biopsies were previously described.^13^ Briefly, 19 adult patients with clinically and histopathologically verified DKD were enrolled at two centers in Sweden between 2010 and 2016. The kidney biopsy tissues, sampled at time of diagnosis, were microdissected into glomerular and tubulointerstitial fractions and profiled with high-throughput RNA sequencing. Written informed consent was obtained from all patients at study enrollment, and the study was approved by Swedish Ethical Review Authorities at both sites (Gothenburg and Stockholm) in accordance with the Declaration of Helsinki 1975, as revised in 2013.

In this study, the patients were followed clinically up to five years post-biopsy and kidney function and events were extracted from patients’ files. eGFR was calculated using the CKD-EPI formula but without modifying for race.^14^ All patients were on standard of care treatment.

### Disease progression definition and cohort stratification

For patients with at least two time-points, “rapid progressor” status was defined according to either one of the following criteria:

1. Negative **eGFR slope** greater than -5 ml/min/1.73m^2^/year;
2. Advancement of the **CKD Stage** category compared to baseline^19^;
3. ≥30% increase in Urine Albumin-Creatinine Ratio (**UACR**) compared to baseline levels^20^;
4. Nephrotic-range albuminuria (**NRA**, defined as UACR > 220mg/mmol) at the last recorded time-point^19^;
5. **Composite outcome** of kidney failure (KF, defined as the initiation of kidney replacement therapy) or ≥40% eGFR decline. ^21–24^

### Kidney transcriptomics analysis in association with DKD progression

Differential expression analysis (DEA) comparing the kidney transcriptomes of rapid progressors and non-rapid progressors was performed using DESeq2 v1.34.0.^25^ Research center factor and relevant covariates that are known to affect disease progression (age, sex, BMI, eGFR and UACR at baseline)^26^ were included in the model design. The p-values were adjusted for multiple testing using the Benjamini-Hochberg method. Genes with an adjusted *P* < 0.05 were considered differentially expressed.

Gene functional annotation was performed using IPA: Ingenuity Pathway Analysis (QIAGEN Inc., https://www.qiagen.com/us/products/discovery-and-translational-research/next-generation-sequencing/informatics-and-data/interpretation-content-databases/ingenuity-pathway-analysis) to determine protein class, subcellular location, upstream regulators, and biomarker properties.^27^ DAVID 2021 was used for Gene Ontology annotation^28–31^ and GTEx v8 for kidney cortex enrichment via z-score calculation.^32^ Weighted Gene Co-expression Network Analysis (WGCNA)^33^ was performed and then visualized using Cytoscape.^34^ Case-control modulation of the progression genes in previous DKD transcriptomics studies (GSE30122, GSE142025, GSE104948, and GSE104954) were also obtained through DEA.^11–12,35–36^

Over-representation analysis was performed using the Reactome database version 82 and Reactome Pathway Browser v3.7,^37^ as well as DAVID 2021. Gene set enrichment analysis was performed with the GSEAPreranked function of GSEA v4.3.2 using Gene Ontology gene sets v2022.1.^30–31,38^

### Statistical analyses

All statistical analyses were conducted using R version 4.1.3.^15^ Clinical parameters were checked for normality of distributions, intercorrelations and lack of center bias. eGFR slopes were estimated using a mixed model repeated measures analysis, with eGFR as the dependent variable, time/visit number and baseline eGFR as linear covariates, and patient as a random effect.^16,17^ Transcriptomics data were filtered and only robustly expressed genes (TPM ≥ 1^18^ in all patients) were included in the downstream analyses: 10 074 genes in glomeruli and 11 964 genes in tubulointerstitium.

## RESULTS

### Characterization of progressive kidney function loss in the study population

Clinical follow-up data were available for 18 out of 19 patients. Individual and categorized eGFR and UACR trajectories are presented in **Figures 1-5**. During the median follow-up of three years (max five years), 13 patients reached the composite endpoint defined as KF or at least 40% eGFR decline. A total of 17 patients had at least two eGFR time-points and the calculated eGFR slopes ranged from -14 to 3.6 ml/min/year. Ten patients had eGFR declining more than -5 ml/min/year, while 11 patients progressed to a more advanced CKD stage. Out of the 16 patients with at least two measurements of UACR, six patients had at least a 30% increase in UACR and nine patients had NRA during their last clinical follow-up.

**Figure 1.**
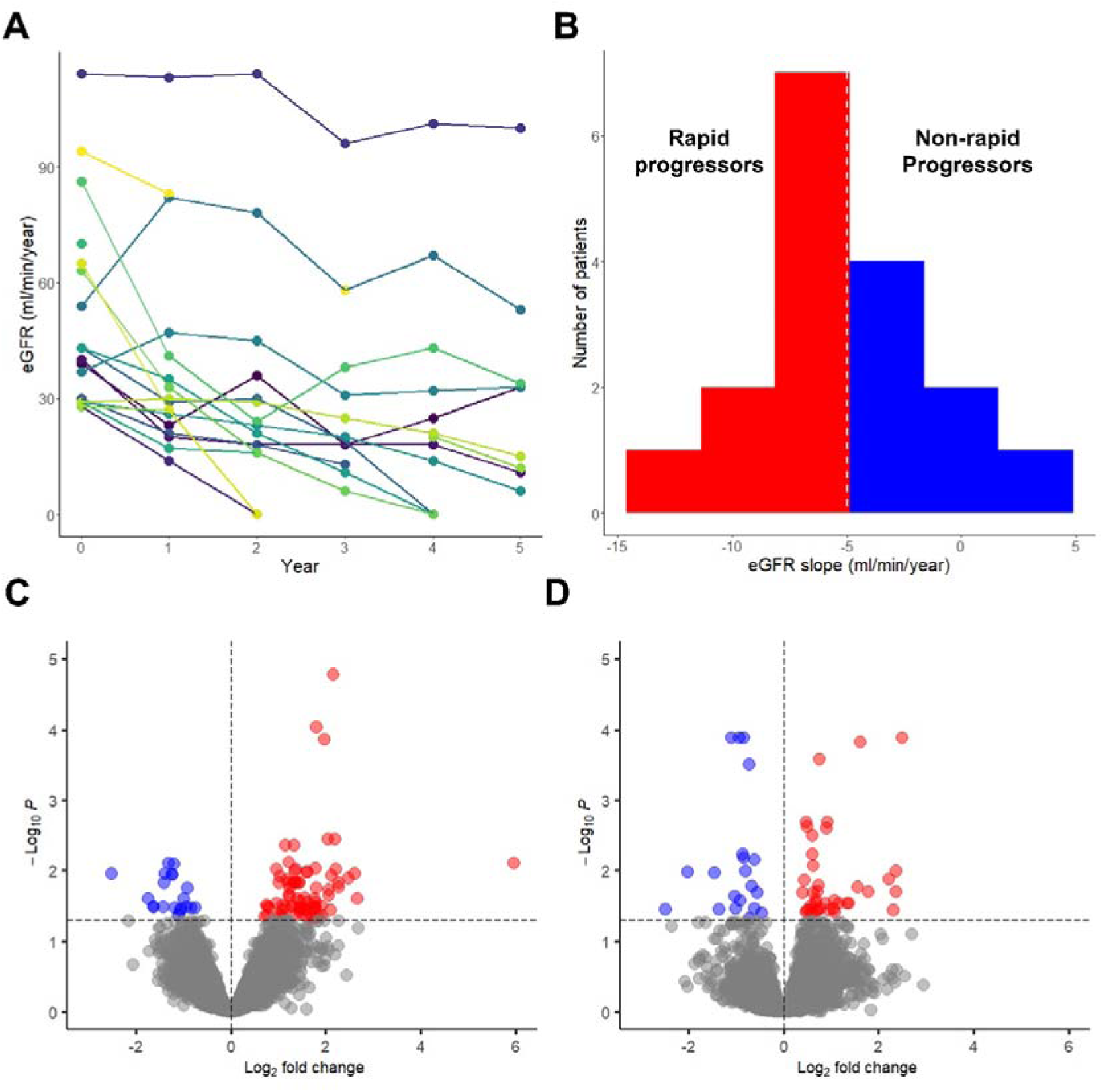
Rapid progression analysis by steep eGFR slope. (A) Individual eGFR trajectories during follow-up. The time-series plot present individual eGFR values from baseline (Year 0) through the duration of follow-up (Year 1-5). eGFR values were set to 0 for patients that reached kidney failure. (B) Histogram of calculated eGFR slopes distributions. Non-rapid progressors are in blue (eGFR slope > -5 ml/min/ year) while rapid progressors are in red (eGFR slope < -5 ml/min/year). (C) DEA of rapid vs non-rapid progressor status in glomeruli. (D) DEA of rapid vs non-rapid progressor status in tubulointerstitium. (C,D) Volcano plots show the log_2_ fold changes in gene expression (x-axis) and the negative logarithm of the adjusted p-value (y-axis) between the rapid and non-rapid progressors. The horizontal dashed line indicates the significance threshold at adjusted P < 0.05 while the vertical dashed line indicates zero fold change. Each point above the horizontal line corresponds to a significantly differentially expressed gene (red: up-regulated, blue: down-regulated). DEA, differential expression analysis; eGFR, estimated glomerular filtration rate.

**Figure 2.**
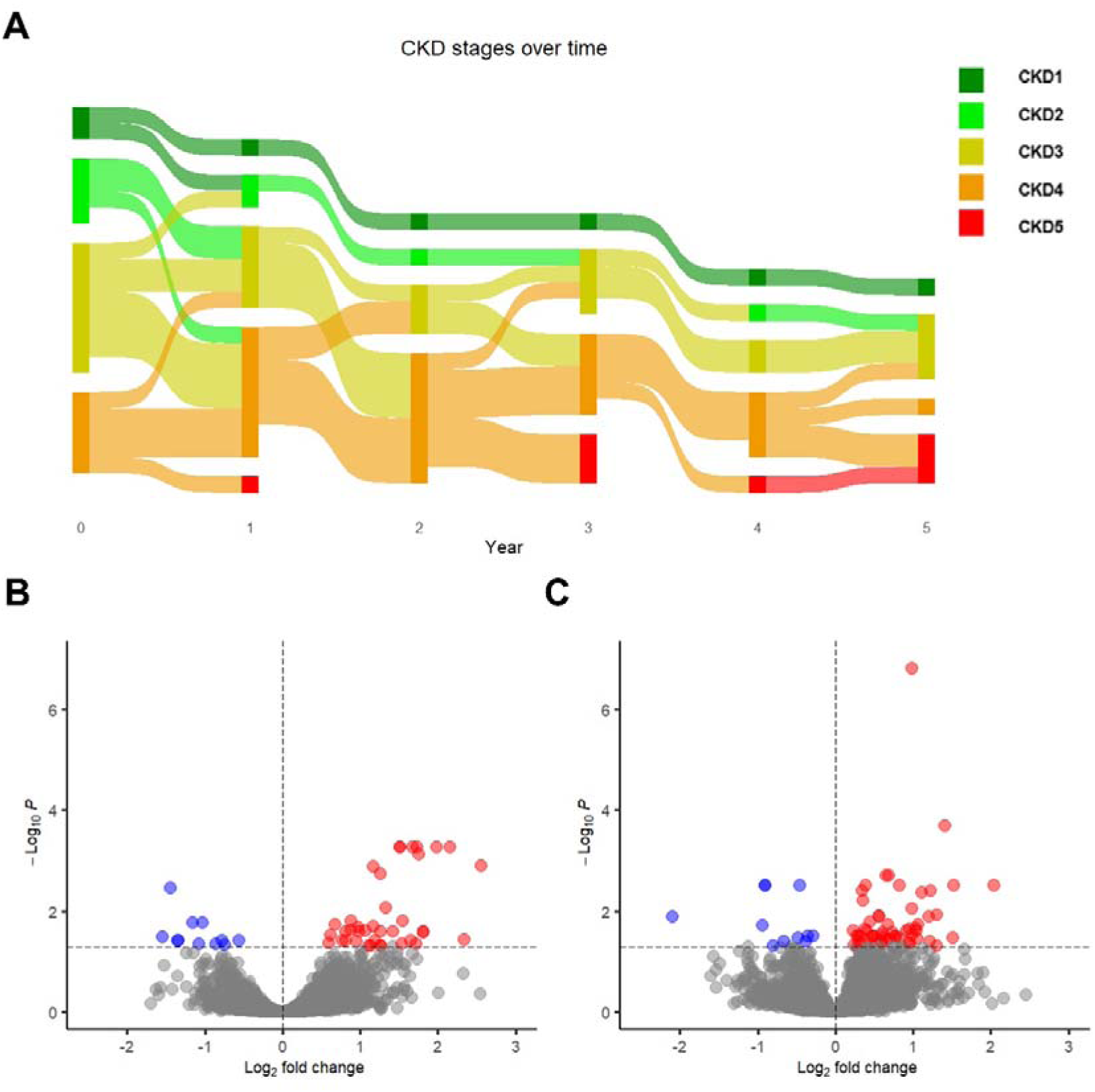
Rapid progression analysis by CKD stage advancement beyond baseline. (A) CKD stage classification changes during follow-up. The Sankey plot demonstrate changes in CKD stage from baseline (Year 0) through the duration of follow-up (Year 1-5). The color ribbons reflect the patient subgroup sizes per category. The unequal number of patients per timepoint exemplifies patients lost to follow-up. (B) DEA rapid vs non-rapid progressor status in glomeruli. (C) DEA rapid vs non-rapid progressor status in tubulointerstitium. (B,C) Volcano plots show the log_2_ fold changes in gene expression (x-axis) and the negative logarithm of the adjusted p-value (y-axis) between the rapid and non-rapid progressors. The horizontal dashed line indicates the significance threshold at adjusted P < 0.05 while the vertical dashed line indicates zero fold change. Each point above the horizontal line corresponds to a significantly differentially expressed gene (red: up-regulated, blue: down-regulated). CKD, chronic kidney disease; DEA, differential expression analysis; eGFR, estimated glomerular filtration rate.

**Figure 3.**
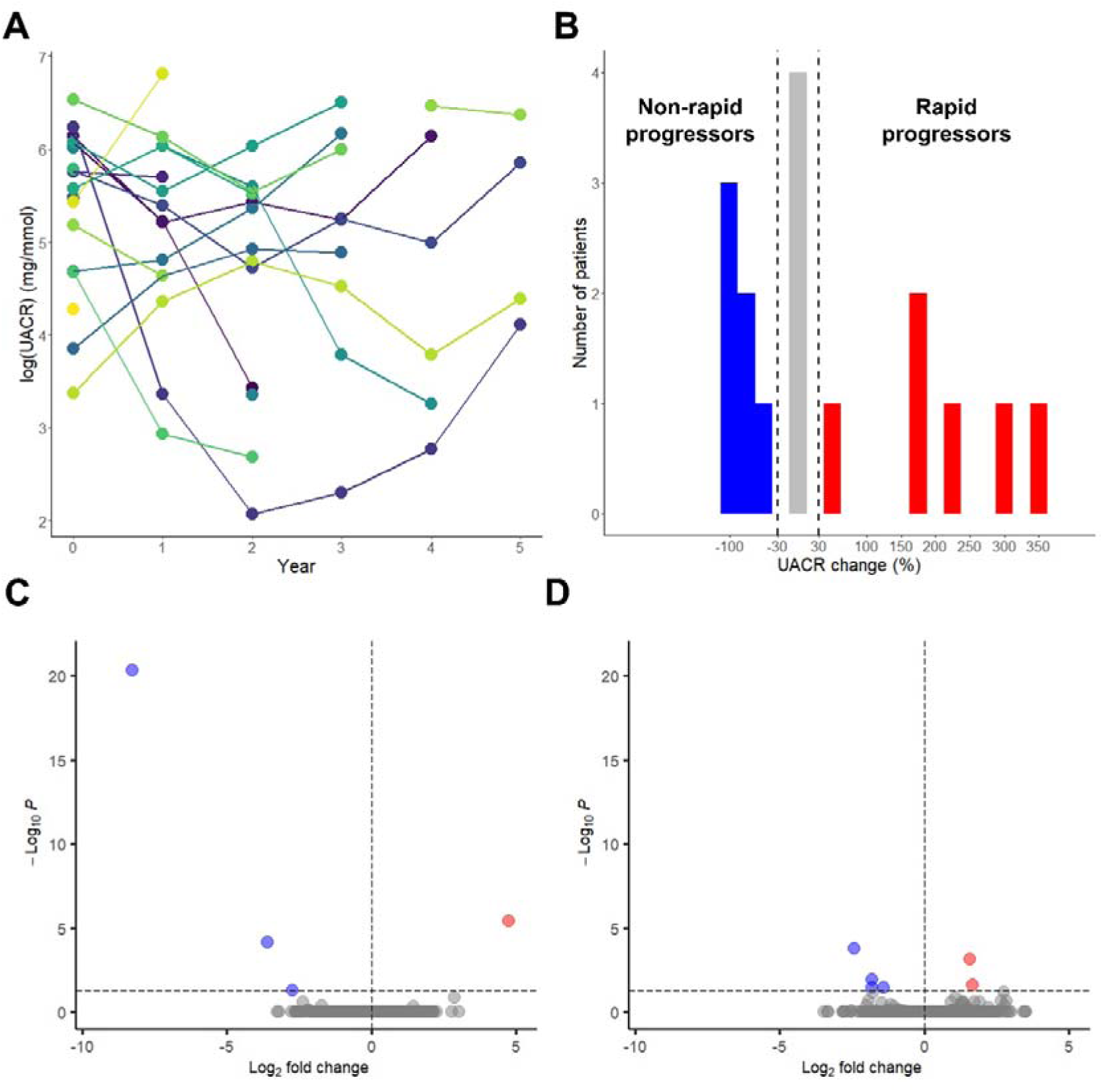
Rapid progression analysis by percentage of UACR change. (A) Individual UACR trajectories during follow-up. The time-series plot present individual log(UACR) values from baseline (Year 0) through the duration of follow-up (Year 1-5). (B) Histogram of percentage of UACR change distributions. Non-rapid progressors are in blue (≥30% UACR decrease) while rapid progressors are in red (≥30% UACR increase). (C) DEA of rapid vs non-rapid progressor status in glomeruli. (D) DEA of rapid vs non-rapid progressor status in tubulointerstitium. (C,D) Volcano plots show the log_2_ fold changes in gene expression (x-axis) and the negative logarithm of the adjusted p-value (y-axis) between the rapid and non-rapid progressors. The horizontal dashed line indicates the significance threshold at adjusted P < 0.05 while the vertical dashed line indicates zero fold change. Each point above the horizontal line corresponds to a significantly differentially expressed gene (red: up-regulated, blue: down-regulated). DEA, differential expression analysis; eGFR, estimated glomerular filtration rate; UACR, urinary albumin-to-creatinine ratio.

**Figure 4.**
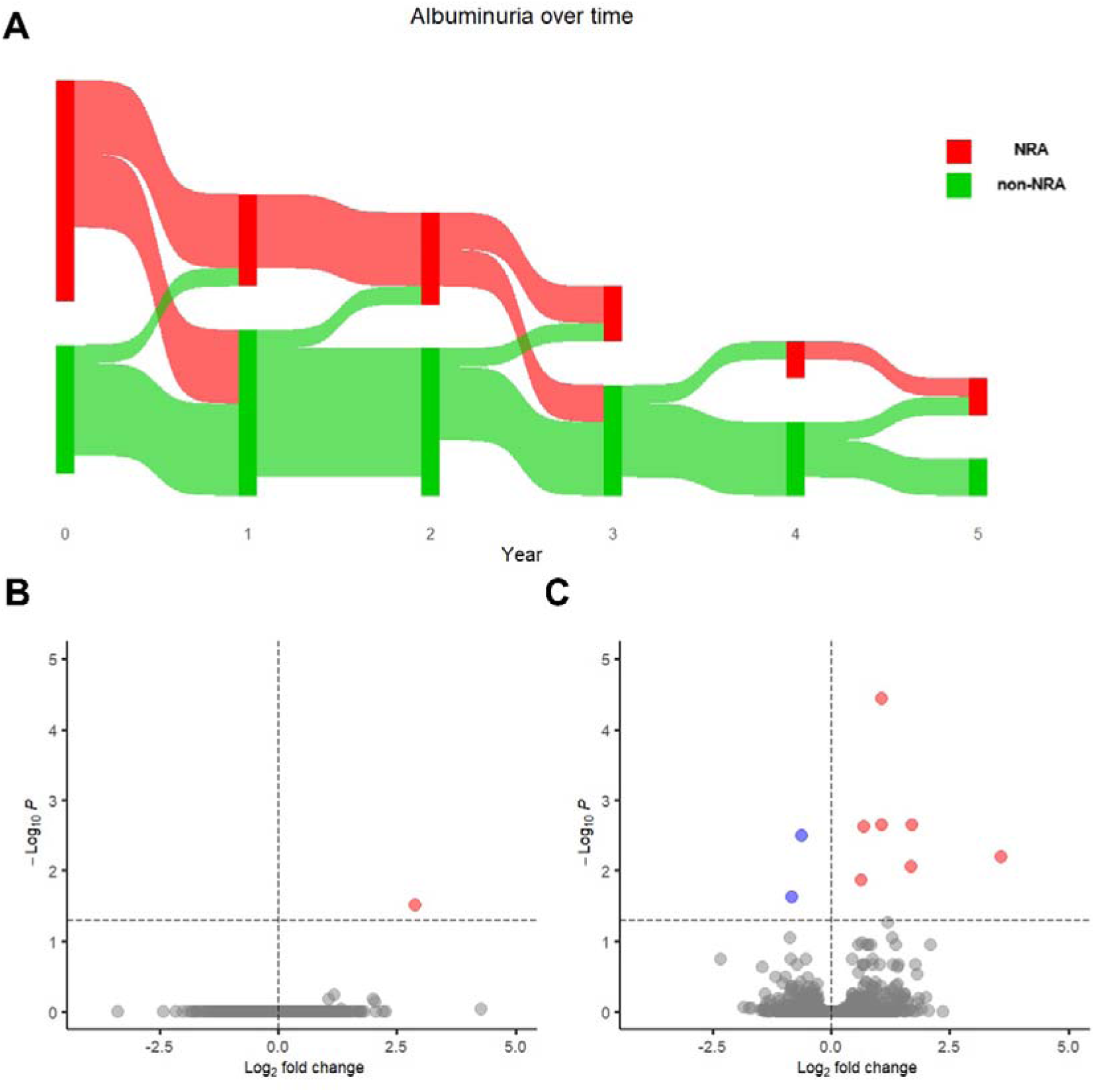
Rapid progression analysis by nephrotic-range albuminuria status. (A) NRA classification changes during follow-up. The Sankey plot demonstrate changes in NRA status from baseline (Year 0) through the duration of follow-up (Year 1-5). The color ribbons reflect the patient subgroup sizes per category. The unequal number of patients per timepoint exemplifies patients lost to follow-up. (B) DEA rapid vs non-rapid progressor status in glomeruli. (C) DEA rapid vs non-rapid progressor status in tubulointerstitium. (B,C) Volcano plots show the log_2_ fold changes in gene expression (x-axis) and the negative logarithm of the adjusted p-value (y-axis) between the rapid and non-rapid progressors. The horizontal dashed line indicates the significance threshold at adjusted P < 0.05 while the vertical dashed line indicates zero fold change. Each point above the horizontal line corresponds to a significantly differentially expressed gene (red: up-regulated, blue: down-regulated). CKD, chronic kidney disease; DEA, differential expression analysis; eGFR, estimated glomerular filtration rate; NRA, nephrotic-range albuminuria.

**Figure 5.**
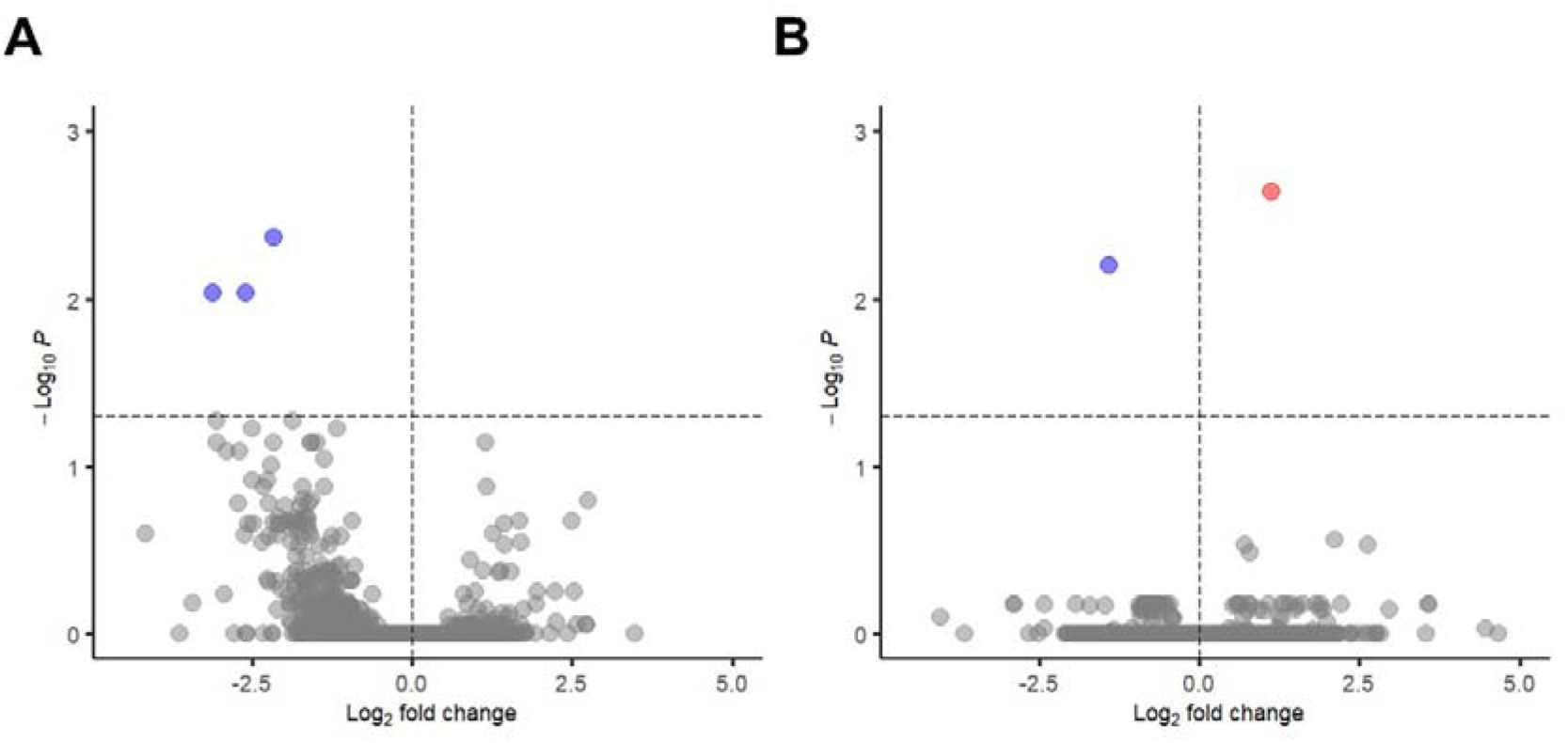
Rapid progression analysis by composite outcome. (A) DEA rapid vs non-rapid progressor status in glomeruli. (B) DEA rapid vs non-rapid progressor status in tubulointerstitium. (A,B) Volcano plots show the log_2_ fold changes in gene expression (x-axis) and the negative logarithm of the adjusted p-value (y-axis) between the rapid and non-rapid progressors. The horizontal dashed line indicates the significance threshold at adjusted P < 0.05 while the vertical dashed line indicates zero fold change. Each point above the horizontal line corresponds to a significantly differentially expressed gene (red: up-regulated, blue: down-regulated). DEA, differential expression analysis.

### Kidney gene expression signature of rapid DKD progression

DEA contrasting rapid progressors versus non-rapid progressors (according to each of the five DKD progression definitions) revealed statistically significant differences in both glomerular and tubulointerstitial transcriptomes, adjusting for known clinical predictors (**Figures 1-5**, **Table 1**). The full DEA output is shared in **Supplementary Data File S1**.

**Table 1.** Counts of significantly modulated genes in the kidneys of rapid progressors vs non-rapid progressors.

| **Rapid disease progression definition** | **Kidney tissue compartment** | **Total number of differentially expressed genes** | **Number of Up-regulated genes** | **Number of Down-regulated genes** |
| --- | --- | --- | --- | --- |
| **eGFR slope < -5ml/min/year** | Glomerular | 97 | 77 | 20 |
|  | Tubulointerstitial | 59 | 39 | 20 |
| **CKD stage advancement** | Glomerular | 49 | 38 | 11 |
|  | Tubulointerstitial | 66 | 55 | 11 |
| **≥ 30% UACR increase** | Glomerular | 4 | 1 | 3 |
|  | Tubulointerstitial | 6 | 2 | 4 |
| **NRA status** | Glomerular | 1 | 1 | 0 |
|  | Tubulointerstitial | 9 | 7 | 2 |
| **Composite of KF or ≥40% eGFR decline** | Glomerular | 3 | 0 | 3 |
|  | Tubulointerstitial | 2 | 1 | 1 |

Rapid progression criteria based on the steep eGFR slope and CKD stage advancement yielded the highest numbers of differentially expressed genes (**Table 1**). In total, we identified 265 rapid progression-associated genes—141 genes in the glomeruli and 134 genes in the tubulointerstitium (**Supplementary Data File S2**), with a modest overlap of 10 genes (*MDK*, *TNFAIP2*, *LINC00152*, *C2orf40*, *ARRB2*, *MIR4435-1HG*, *MMP7*, *AL353644.10*, *COL8A1*, *MTND2P28*) shared between the glomerular and tubulointerstitial fractions.

### In silico characterization of the rapid DKD progression signature

To further characterize the rapid progression-associated genes, they were annotated with functional properties and investigated in terms of enriched biological pathways, co-expression, transcriptional regulators, and associations in previous case-control studies (**Supplementary Data File S2**).

The rapid progression-associated genes were diverse in terms of their protein products’ classes, which included cytokines, enzymes, G-protein coupled receptors, growth factors, ion channels, kinases, peptidases, phosphatases, transcriptional and translational regulators, transmembrane receptors, and transporters. They displayed predominantly cytoplasmic subcellular localization (103), followed by plasma membrane (51), extracellular space (44), and nucleus (29). Several genes in the rapid progression signature showed strong preferential expression in the healthy kidney cortex as compared to other tissues, such as *AQP6*, *CDH16*, *DHDH*, *LHFPL3-AS2*, *LINC01055*, *LINC01671*, *REN*, *SLC12A3*, and *SPP1*. Based on the IPA BiomarkerFilter analysis, protein products of 124 genes were reported to be measurable in blood, plasma or urine, of which 80 were in the context of urological and nephrological disease, thus supporting their potential biomarker properties.

Upon the compartment-stratified over-representation analysis, the rapid progression-associated genes showed enrichment for specific biological processes (**Figure 6A-D**). Top enriched pathways include integrin cell surface interactions, extracellular matrix assembly and degradation, and interleukin signaling (glomerular subsignature), and pathways for nonsense-mediated decay, ribosomal formation, translation processes, and cellular response to starvation (tubulointerstitial subsignature). Gene set enrichment analysis (GSEA) revealed additional pathways represented in the rapid progression signature such as mitochondrial dysfunction, oxidative stress, ubiquitination and circadian rhythm regulation. The complete pathway enrichment analyses results are provided in **Supplementary Data Files S3 and S4**.

**Figure 6.**
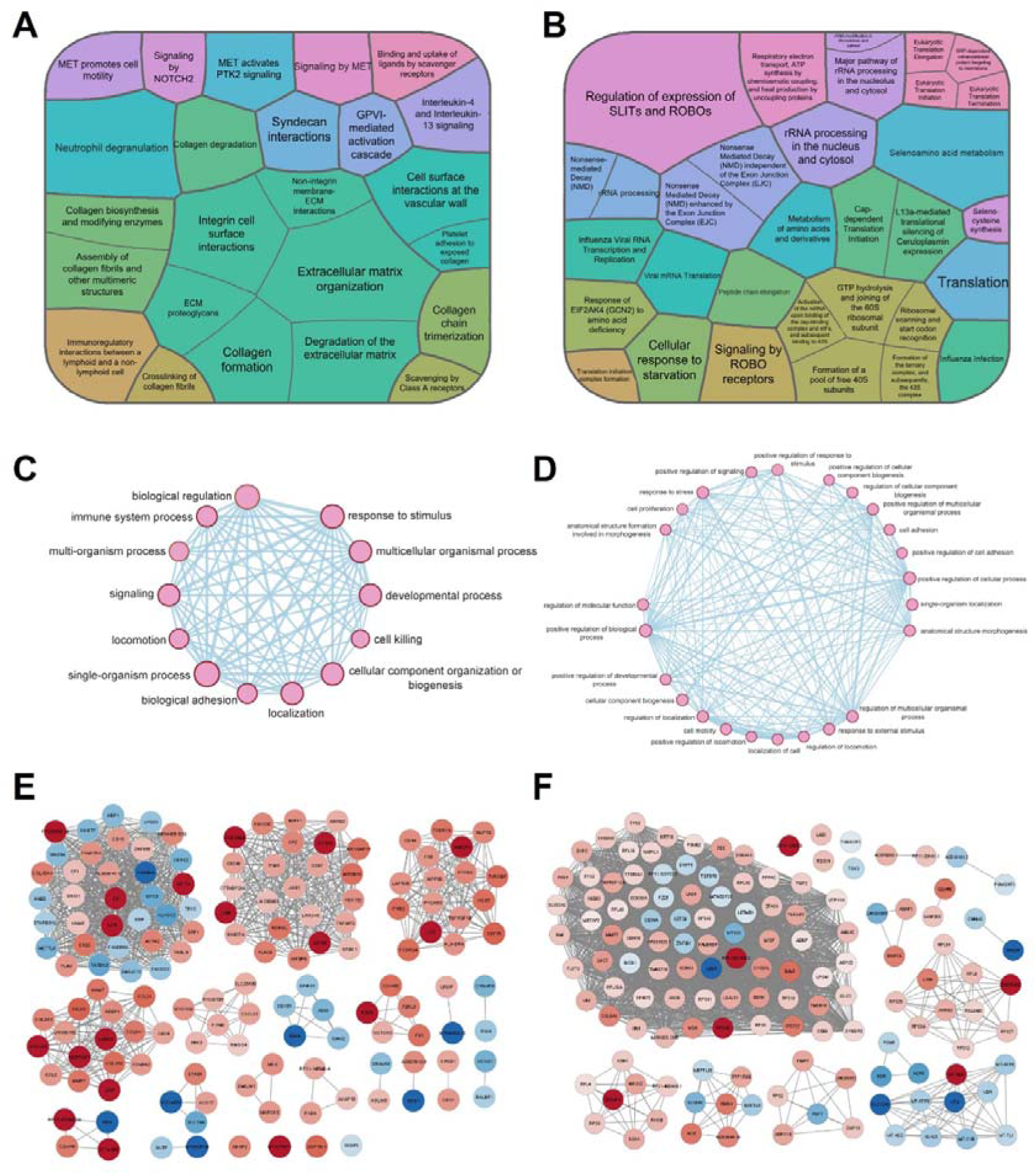
Rapid progression genes and associated pathways. (A,B) Treemaps show the results of the Reactome pathway over-representation analysis of rapid progression-associated genes according to all five disease progression definitions in the (A) glomeruli and (B) tubulointerstitium. The color of the cells denotes biologically related pathways (same Reactome parent pathway). The size of the cells reflects the size of the overlap between the progression-associated genes and the corresponding pathway. Only significant terms (adjusted P < 0.05) are shown. (C,D) Network maps show the significant (FDR < 0.05) Gene Ontology biological processes in the (C) glomeruli and (D) tubulointerstitium determined through over-representation analysis. (E,F) Co-expression network maps of the rapid progression-associated genes in the (E) glomeruli and (F) tubulointerstitium. Connected genes are co-expressed in the specified module or subnetwork. Genes are colored based on log_2_ fold changes, where red and blue indicate up-regulation and down-regulation, respectively. FDR, false discovery rate.

The upstream regulator analysis revealed that some of the rapid-progression-associated genes share common transcriptional factors, including *MAFB*, *STAT3*, *SMARCA4*, *HIF1A*, *TP63*, *JUN*, *YAP1*, and *GLI1* in the glomeruli and *SREBF1*, *GLI1*, *TP73*, *YAP1*, *MYC*, *SMARCA4*, and *SP1* in the tubulointerstitium. The comprehensive list of upstream regulators are shared in **Supplementary Data File S5.**

In line with shared transcriptional regulation, the co-expression analysis revealed subnetworks among the rapid progression genes based on gene correlation and module membership (**Figure 6E-F**). Screening for highly connected genes in each module showed that *C16orf58*, *COL1A1*, *CXCL12*, *EMILIN1*, *GLIPR2*, *ITGB2*, *NNMT* and *TNFRSF1B* are hub genes in the glomeruli and *RHOB* in the tubulointerstitium.

Looking over four publicly available DKD data sets, 39 of the 265 rapid progression-associated genes have not been previously reported as differentially expressed between diseased vs healthy patients. The majority of the progression genes (160) were significantly modulated in the same direction in rapid vs non-rapid progressors as in DKD cases vs controls. A small percentage of the progression transcripts either have conflicting modulation directionality (47) or have not been sequenced in previous cross-sectional DKD transcriptomics studies (19). A summary is presented in **Supplementary Data File S6**.

## DISCUSSION

To the best of our knowledge, this is the first longitudinal study of DKD kidney biopsy transcriptomics with prospectively collected clinical outcomes data. Such a design has the potential to advance knowledge beyond the amassed cross-sectional evidence and gain novel insights into the local molecular mechanisms driving the disease progression, thus contributing value to the field with respect to deepening biological understanding and ultimately biomarker and drug development.

We identified in total 265 intra-renal transcripts associated with rapid progression of DKD based on different clinical outcome definitions in a model adjusted for baseline kidney function. The majority of the progression-associated genes were up-regulated, which likely reflect increased transcriptional activity, whereas the interpretation of the down-regulated genes may be obscured due to loss of resident cells in the diseased kidney.

Of note, the different clinical phenotypes of rapid progression highlighted diverse gene expression patterns, thus capturing distinct biological nuances of progressive DKD. Stratification using eGFR slope and CKD stage advancement generated the highest numbers of differentially expressed genes. On the other hand, albuminuria-based analyses were hampered by the lack of UACR variability in this high-risk near-uniformly macroalbuminuric population and, therefore, produced few significant hits.

Interestingly, the DKD progression-associated transcriptomics signature displayed a high degree of kidney compartment specificity, which is in line with biological expectations based on the function of different kidney cell types and independent contributions of glomerular and tubular pathologies to kidney outcomes.^39^ Furthermore, the progression-associated genes are likely not independent players as they displayed co-expression and shared transcriptional regulation, implying participation in common broader biological programs.

This indicates the possibility to select “hub” targets for intervention in order to develop new therapeutic modalities with pleiotropic beneficial effects.

As the heterogeneity of the clinical course of DKD patients remains a challenge, there is a strong need for novel non-invasive biomarkers to enable early prediction of rapid progression to optimally allocate patient care and increase the likelihood of slowing down the kidney function loss in high-risk patients. Almost 50% of the protein products associated with rapid progression in our study are excreted and measurable in biofluids. These potential biomarkers, if validated in large cohort studies and implemented in clinical testing, could improve prediction of DKD progression and would add valuable information besides traditional clinical risk factors.

The most prominent biological themes reflected in the rapid progression signature included many known disease-associated pathways, thus corroborating the domain knowledge. The fact that several rapid progression-associated genes also showed modulation in previous cross-sectional studies add evidence of them having either a potential causal role in driving or perpetuating pathophysiological processes. However, a number of progression-associated genes did not show case-control modulation and thus would have been missed previously. Interestingly, several progression-associated genes are notable to have strong enrichment compared to other organs under normal conditions, thus supporting their kidney relevance and potentially suggesting that they play a role both under healthy and pathophysiological conditions.

For example, the top significant progression-associated transcript in the glomerular subsignature was the kidney-enriched *REN* which encodes renin and is expressed exclusively by the specialized juxtaglomerular cells.^40^ It was strongly down-regulated in the rapid progression cases developing ≥30% UACR increase post-baseline compared to those who did not. The renin-angiotensin-aldosterone system (RAAS) is a well-validated therapeutic target in DKD and CKD, and RAAS inhibitors (RAASi) remain standard of care in DKD and CKD due to their blood pressure and albuminuria lowering effects, thereby preventing eGFR decline.^41^ Of note, RAASi treatment is known to impact *REN* mRNA levels through transcriptional regulation, which could make interpretation of patient transcriptomics data challenging.^42^ However, all the participants in this study were on RAASi treatment at the time of biopsy.

The top significant gene in the tubulointerstitium was *SOX4*, SRY box transcription factor 4, up-regulated by almost 200% in the rapid progressors whose CKD stage advanced during follow-up. The SOX4 protein is a transcriptional factor involved in several different processes including kidney embryogenesis,^43^ tumorigenesis by regulating cell differentiation,^44^ and apoptosis induction.^45^ It is expressed in fetal human kidneys^46–47^ and in early tubules of mice.^48^ The role of *SOX4* in the adult kidney remains to be elucidated, but since apoptosis and dedifferentiation of proximal tubular epithelial cells are well-known features of DKD,^49–50^ *SOX4* can be hypothesized to be involved.

Additional pathways that were highlighted among the rapid progression-associated genes in the glomeruli were cytoskeletal rearrangement and circadian rhythm. *NOTCH3* was up-regulated by almost 200% in the rapid progressor group with a steep negative eGFR slope. *NOTCH* pathway activation has previously been observed in DKD glomeruli with podocyte loss^51^ and *NOTCH3*, specifically, is implicated in pro-inflammatory and pro-fibrotic changes in the podocytes.^52^ Cytoskeleton reorganization also goes hand in hand with podocyte injury and proteinuria.^5^ Circadian rhythm dysregulation has been linked to decreased GFR^53^ and increased risk of CKD.^54^ The glomerular extracellular matrix is subject to local circadian dynamics, and there is mounting evidence suggesting that disruption of the intra-glomerular cellular clock plays a role in disease pathogenesis.^55^ Taken together, the rapid progression-associated genes and pathways in glomeruli appear to be all in interplay.

In the tubulointerstitial signature, mitochondrial and endoplasmic reticulum (ER) processes emerged. Mitochondrial oxidative phosphorylation is the main source of ATP-based energy in the kidney. In DKD, a decrease in the mitochondrial membrane potential in the proximal tubules initiates mitophagy leading to mitochondrial dysfunction and the metabolic switching to glycolysis to compensate for unmet ATP demands.^56^ Consistent with this, we found the mitochondrial oxidative phosphorylation pathways were transcriptionally inhibited in the tubulointerstitium of rapidly progressing DKD cases, likely reflecting impaired metabolism or failure of compensatory mechanisms.

*MAP2K3* was strongly up-regulated in the tubulointerstitium of rapid progression cases at risk for developing nephrotic-range albuminuria. *MAPK* signaling can be modulated in response to ER stress^57^ and its activation has been shown to be implicated in tubulointerstitial fibrosis.^58^ Genetically *MAP2K3*-deficient diabetic mice had reduced *MAPK* signaling and observed to be protected from declining kidney function and increasing albuminuria.^59^

Finally, ribosomal translation processes were activated in the tubulointerstitium of rapid progressors based on steep negative eGFR decline and CKD stage advancement. Translation pathways have been reported enriched in the proximal tubules of DKD kidney biopsies compared to controls.^60^ This increase in mRNA translation could be caused by matrix accumulation and higher amounts of growth factors, both of which are prevalent in tubular injury and progressive DKD.^61^ An imbalance brought about by an increase in protein synthesis and degradation can progress to ER stress,^60^ which is one of the critical mechanisms in albumin-induced tubular injury.^62^

Several limitations of this study should be acknowledged. First, the sample size was limited, even though this DKD dataset remains the largest available with kidney biopsy compartment-specific RNA-seq transcriptomics profiling. However, despite the modest sample size, there was a sufficient degree of inter-individual heterogeneity and disparate disease progression rates, enabling us to stratify the cohort and identify significant progression-associated transcriptional changes. Larger studies in independent cohorts are warranted to validate these findings and further refine DKD progression mechanistic understanding. Secondly, selection bias cannot be ruled out since a clinically indicated kidney biopsy is rarely performed in DKD and thus, the biopsy cohort may not be representative of the general DKD. However, this study highlighted potential non-invasive biomarker candidates that can be further tested in broader DKD populations. Finally, the duration of the follow-up in this study was limited to five years; it is conceivable that longer clinical observation time might reveal additional biological pathways involved in more chronic processes.

To conclude, our study delivered the first longitudinal analysis linking baseline kidney transcriptomics profile with prospective clinical outcome in diabetic kidney disease. These findings shed light onto the molecular mechanisms driving progressive kidney function decline in DKD, the most common etiology of CKD, and hold the potential to contribute to future diagnostic and therapeutic development.

## Data Availability

The raw RNA-seq data analyzed in this study is openly available at http://karokidney.org/rna-seq-dn/.

## ACKNOWLEDGMENTS

This research was supported by grants provided by the Stockholm County Council (ALF Project), and the European Union Horizon 2020 research and innovation programme under the Marie Skłodowska-Curie grant agreement No. 860977 titled TrainCKDis.

Initial preliminary results from this work were previously shared as an abstract and poster at the World Congress of Nephrology 2023 (March 30 - April 2, 2023) and as an abstract and oral presentation at the ASN Kidney Week 2023 (November 2-5, 2023).

## AUTHOR CONTRIBUTIONS

All co-authors have contributed to the manuscript. Specifically, A.We., A.Wi, J.N, P.J.G., A.R. designed and directed the project. A.L. collected the data. D.A. analyzed the data and produced the figures. All authors interpreted the results, and provided critical feedback that helped shape the research, analysis and manuscript. D.A., A.L., A.Wi. drafted the manuscript. All authors approved the final version and accept accountability for the overall work by ensuring that questions pertaining to the accuracy or integrity of any portion of the work were appropriately investigated and resolved.

